# Genomic Surveillance for Enhanced Healthcare Outbreak Detection and Control

**DOI:** 10.1101/2024.09.19.24313985

**Authors:** Alexander J. Sundermann, Praveen Kumar, Marissa P. Griffith, Kady D. Waggle, Vatsala Rangachar Srinivasa, Nathan Raabe, Emma G. Mills, Hunter Coyle, Deena Ereifej, Hannah M. Creager, Ashley Ayres, Daria Van Tyne, Lora Lee Pless, Graham M. Snyder, Mark Roberts, Lee H. Harrison

## Abstract

**Background:** Current methods are insufficient alone for outbreak detection in hospitals. Real-time genomic surveillance using offers the potential to detect otherwise unidentified outbreaks. We initiated and evaluated the Enhanced Detection System for Healthcare-associated Transmission (EDS-HAT), a real-time genomic surveillance program for outbreak detection and mitigation.

**Methods:** This study was conducted at UPMC Presbyterian Hospital from November 2021 to October 2023. Whole genome sequencing (WGS) was performed weekly on healthcare-associated clinical bacterial isolates to identify otherwise undetected outbreaks. Interventions were implemented in real-time based on identified transmission. A clinical and economic impact analysis was conducted to estimate infections averted and net cost savings.

**Results:** There were 3,921 bacterial isolates from patient healthcare-associated infections that underwent WGS, of which 476 (12.1%) clustered into 172 outbreaks (size range 2-16 patients). Of the outbreak isolates, 292 (61.3%) had an identified epidemiological link. Among the outbreaks with interventions, 95.6% showed no further transmission on the intervened transmission route. The impact analysis estimated that, over the two-year period, 62 infections were averted, with gross cost savings of $1,011,146, and net savings of $695,706, which translates to a 3.2-fold return on investment. Probabilistic sensitivity analysis showed EDS-HAT was cost-saving and more effective in 98% of simulations.

**Conclusion:** Real-time genomic surveillance enabled the rapid detection and control of outbreaks in our hospital and resulted in economic benefits and improvement in patient safety. This study demonstrates the feasibility and effectiveness of integrating genomic surveillance into routine infection prevention practice, offering a paradigm shift in healthcare outbreak detection and control.

## INTRODUCTION

Outbreak detection in healthcare settings is a vital aspect of infection prevention because it directs interventions to prevent additional pathogen spread. Despite advances in healthcare technologies, the methods used for outbreak detection have remained largely unchanged for decades.^1^ Traditional methods usually rely on surveillance and monitoring for an observed increased incidence of infections from a baseline level which prompts follow-up investigation.^2^ Whole genome sequencing (WGS) is the current standard for genetic relatedness testing, and when used for this purpose is referred to as “reactive WGS”. However, this approach lacks timeliness, often misidentifies outbreaks that are not confirmed, and misses many, consequential outbreaks altogether.

Genomic surveillance using WGS, or “prospective WGS surveillance” is an emerging approach that overcomes the limitations of current outbreak detection with reactive WGS by enabling the sequencing of pathogens regardless of the presence of an outbreak. Prospective WGS surveillance could identify outbreaks as early as two patients, which would allow for rapid interventions and halt further transmission. This approach in healthcare settings has not been widely adopted for real-time use due to needed investments in WGS infrastructure and lack of incentives.^3^ Nevertheless, recent studies have shown WGS surveillance as a real-time infection prevention and control (IP&C) tool is promising given its potential to prevent infections and avert healthcare costs.^4–6^

We developed the Enhanced Detection System for Healthcare-associated Transmission (EDS-HAT) which uses novel approaches to detect and investigate outbreaks quickly and accurately.^4,7^ After a retrospective analysis showing EDS-HAT’s promise as an effective infection prevention tool, we initiated real-time genomic surveillance in November 2021.^4^ Here, we describe the first two years of findings and implications for both IP&C and cost savings through the prevention of additional infections.

## METHODS

### Study setting

This study was performed at UPMC Presbyterian hospital, an adult tertiary care hospital with 699 total beds, 134 critical care beds, and over 400 annual solid organ transplantations. Ethics approval was obtained from the University of Pittsburgh Institutional Review Board.

### Isolate Collection

Isolate collection occurred twice per week from November 1, 2021 to October 31, 2023. Patients with clinical cultures positive for select bacterial pathogens were included if the patient had been in the hospital for ≥3 days or had a recent UPMC healthcare exposure in the prior 30 days (Supplement Methods). For *Clostridioides difficile*, culture-independent diagnostic test-positive stool specimens were cultured for the organism. Active surveillance cultures were not included.

### Genomic Methods

WGS was performed on the NextSeq 500 platform (Illumina, San Diego, CA). Reads were assembled with Unicycler v0.5, annotated with Prokka v1.14, and multilocus sequence types (STs) were assigned using PubMLST typing schemes (https://github.com/tseemann/mlst). Reads were classified to species level using Kraken2 v2.12.2 with the standard kraken database. Isolates passed quality control if 1) the most prevalent species from Kraken was the expected species, 2) the assembly length was within 20% of the expected genome size, 3) the assembly was ≤350 contigs and 4) there was at least 35X depth. Pairwise single nucleotide polymorphism (SNP) differences were calculated using both SKA v1.0 within all isolates in a species and Snippy v4.3.0 (https://github.com/tseemann/snippy) within species STs having two or more isolates. For each pairwise comparison, the minimum SNP distance between SKA and Snippy was used to calculate outbreaks. Genetically related outbreaks were assigned using initial SNP cutoffs using hierarchical clustering with average linkage. Based on our experience and the literature, an outbreak was defined as isolates from more than one patient having ≤15 pairwise cgSNPs for all species except for *C. difficile*, for which two or fewer pairwise core-genome SNPs (cgSNPs) were used to identify outbreaks.^8^ For a detailed analysis of our SNP threshold, we performed a pairwise SNP analysis of genetically linked isolates to identify epidemiological links for all pathogens, excluding *C. difficile*, using logistic regression analysis (SAS version 9.4, Cary, NC).

### Infection Prevention

Upon detection of an outbreak or a new isolate linked to an outbreak, the research team conducted a thorough review of patient records to identify epidemiological connections, focusing on geo-temporal links, shared procedures, and interactions with common staff members.^4,9^ The initial findings were shared with the IP&C team for a second, in-depth analysis. Based on these findings, the IP&C team promptly initiated further actions including investigations and targeted interventions when indicated. Further investigation may include observation of infection prevention practices (e.g., hand hygiene, personal protective equipment use, cleaning/disinfection) and additional case finding through microbiological surveillance for colonization or infection. Examples of interventions included: staff education, enhanced environmental cleaning, and removing potentially contaminated equipment from service, and other relevant measures. The initiation date of each intervention was recorded by the IP&C team. The IP&C team further noted major changes in practice due to EDS-HAT findings.

### Economic and Clinical Impact Analysis

This analysis focused on evaluating the continuity of transmission on the intervened suspected transmission route. Outbreaks with an identified transmission route were classified as 1) stopped at two cases after IP&C intervention, 2) continued on new transmission route(s) that was not intervened upon (for example, an outbreak is identified on unit A, the intervened upon transmission route, but, because of patient movement, transmission of the outbreak strain occurs on unit B), and 3) continued on the same transmission route despite intervention. The remaining outbreaks were classified as having no identified transmission route.

We used our prior retrospective genomic surveillance data in the setting of no interventions to estimate the impact of our real-time approach and calculated the proportion of outbreaks and average number of patients within an outbreak exceeding two patients stratified by transmission route (e.g., unit-based, endoscope-based).^4^ We applied this proportion of outbreaks to our detected real-time outbreaks and calculated the number of expected patients to exceed the two patient detection threshold. We subtracted the number of observed patients within outbreaks that exceeded two patients by transmission route, indicating an intervention failure (Supplement Methods, Figures S1, S2). This observed minus expected provided the approximate number of infections averted. We incorporated our prior estimates of the cost of treating HAIs and the cost of performing WGS surveillance to calculate the approximate net savings to the hospital.^10,11^ Some outbreaks were detected in non-UPMC Presbyterian Hospital facilities. However, they were not included in the impact analysis because these facilities are not under the control of our IP&C team. Probabilistic sensitivity analysis (PSA) was conducted to assess the assumptions of number of subsequent infections within an outbreak (Supplement Methods, Table S1). All costs were adjusted to 2023 using the medical component of the Consumer Price Index (CPI).^12^

## RESULTS

We identified 7,051 unique isolates of EDS-HAT organisms during the study. Of these, 4,723 (66.9%) were deemed healthcare-associated according to EDS-HAT criteria and underwent WGS. There were 3,921 unique patient infections of which 476 isolates (12.1%) were related to at least one other isolate. Of the outbreak isolates, 419 (88.0%), originated from inpatient samples, while 36 (7.6%) and 21 (4.4%) were collected during emergency room or outpatient encounters, respectively. Average time from culture collection to genomic analysis completion was 15 days (median 14 days, range 7-58 days). There were 172 outbreaks identified, ranging in size from 2 to 16 patients (Table 1, Figure 1, Table S3).

**Figure 1.**
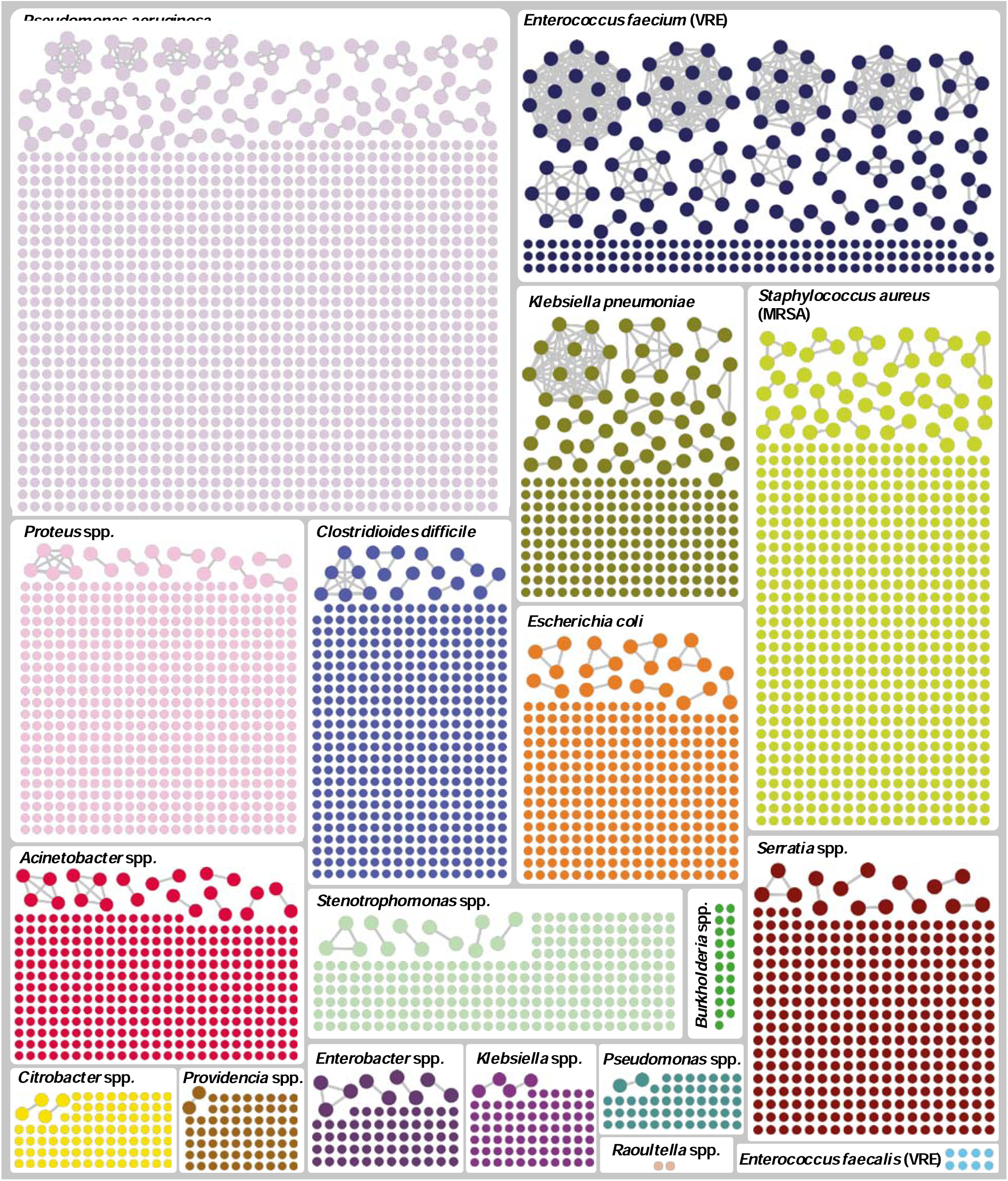
Graphical depiction of isolates that belong to an outbreak (connected dots) versus those that are unrelated (unconnected dots), by species.

**Figure 2.**
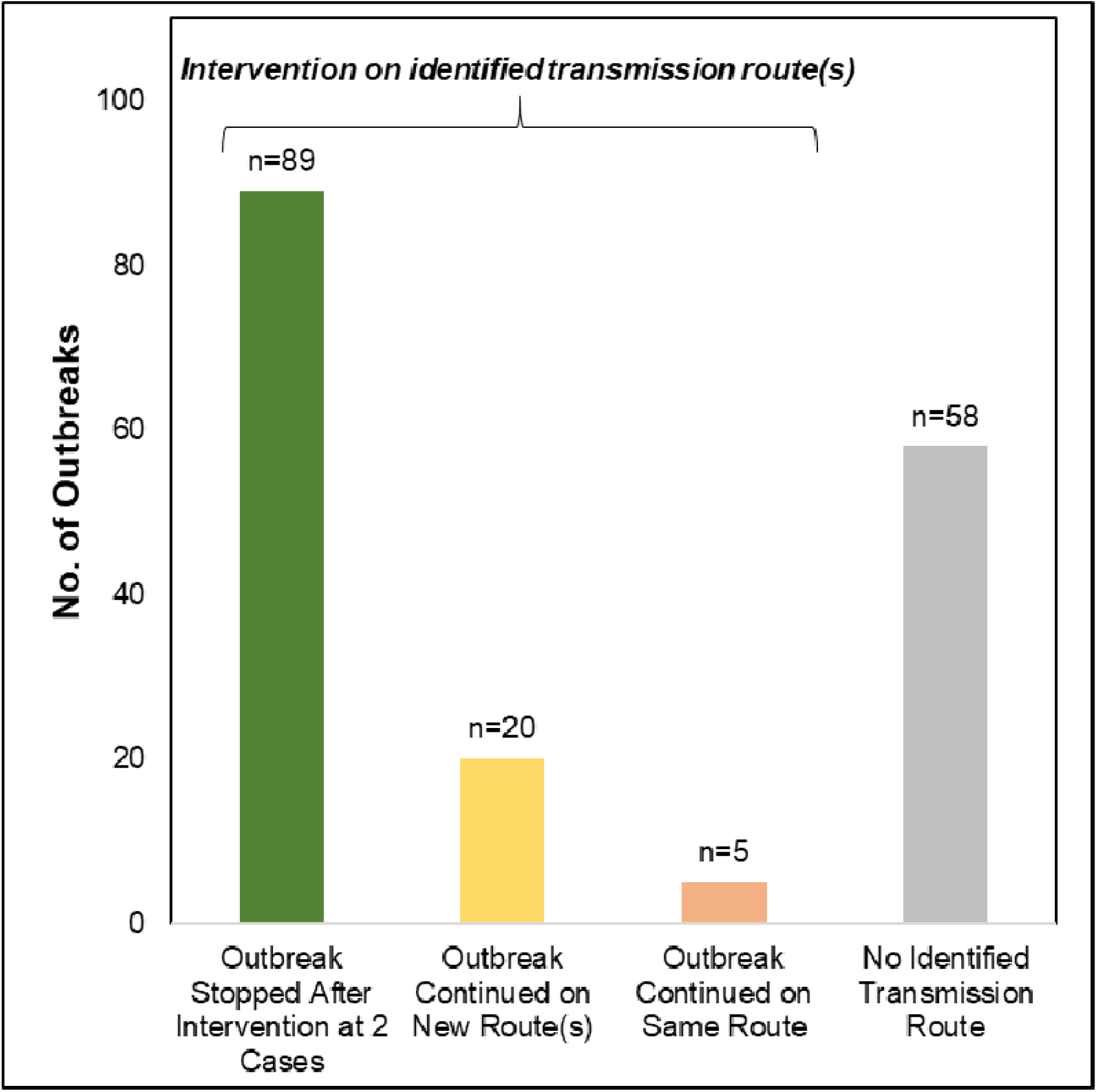
Outbreak outcomes after IP&C intervention, by whether transmission route was identified (first 3 bars) or not (final bar).

**Table 1.**
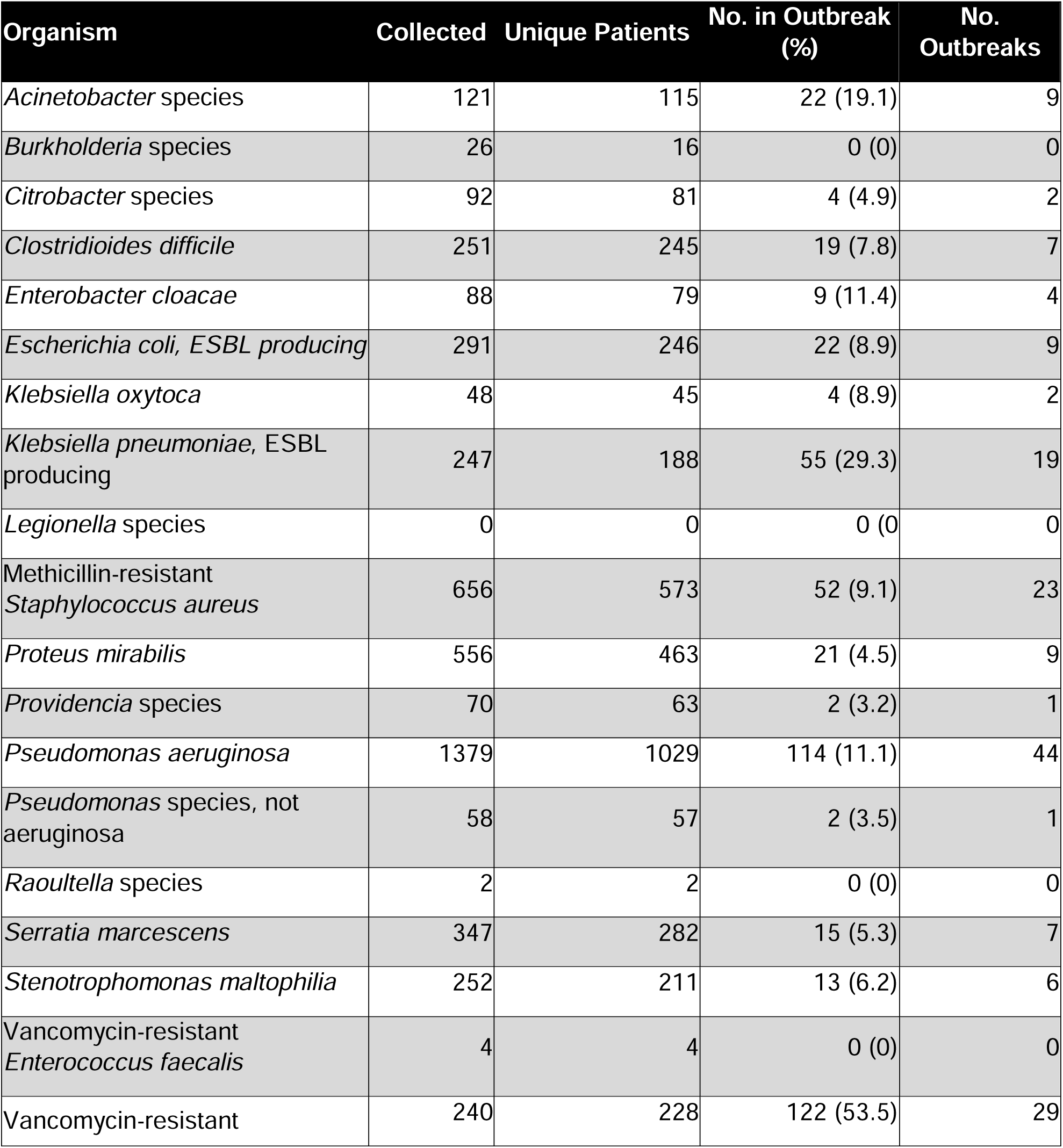
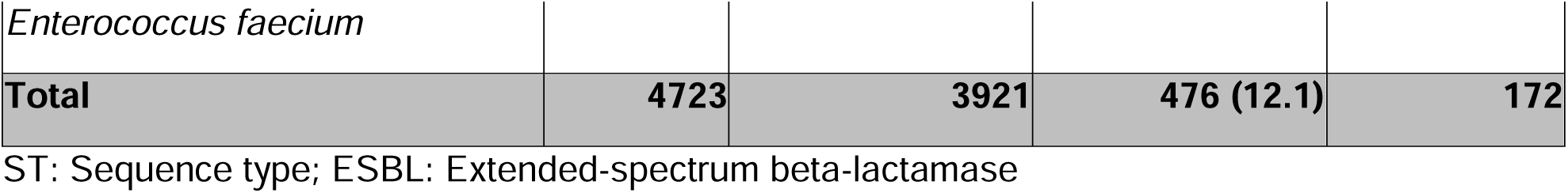
Number (%) of clinical isolates sequenced that were determined to belong to an outbreak (as defined in Methods), by bacterial species, November 2021 – November 2023.

### Presumed transmission routes

Of the 476 outbreak isolates, 292 (61.3%) had an identified epidemiological linkage with another isolate. The most common mode of transmission was unit-based, accounting for 169 isolates (35.5%). Other transmission routes included 39 isolates (8.2%) linked to various sources such as equipment (e.g., ventilators) and healthcare workers, 66 isolates (13.9%) traced back to external healthcare facilities, and 18 isolates (3.8%) associated with shared endoscopes. Notable or high-impact outbreaks are described in Table 2.

**Table 2.** Notable outbreaks, by transmission at UPMC-Presbyterian Hospital versus elsewhere.

| Outbreak | Description |
| --- | --- |
| **Transmission at UPMC Presbyterian Hospital** |  |
| Endoscope-related | A total of nine outbreaks involved 2 patients each with suspected transmission via the same endoscope. Upon detecting each transmission, the implicated endoscopes were immediately removed from service for inspection. Notably, one scope failed a leak test and was never returned to service. Subsequent to the removal of these endoscopes, no additional cases were detected related to the same equipment. Endoscope-related outbreaks that we detected at our institution before we initiated real-time EDS-HAT involved at least 6-17 patients.<sup>13,16</sup> |
| Ventilators | Three outbreaks were identified, involving seven total patients, where in each outbreak, the same ventilators were used sequentially from one patient to the next. In one outbreak, two patients were linked within the same unit; infection in a third patient occurred on a different intensive care unit where the implicated ventilator had been relocated. IP&C educated respiratory care on ventilator cleaning and maintenance practices. No additional cases seen by ventilator transmission. |
| Wound care | A single outbreak involving seven patients over the first year, all linked to vancomycin-resistant *Enterococcus faecium* and potential exposure through wound care services. These patients were dispersed across multiple inpatient units but shared the common exposure of receiving care from the same wound care service. Following targeted education, enhanced observation practices in wound care, and changing of equipment, no additional associated infections were observed. In a prior study, we identified wound care as a possible transmission route for 9 *C. difficile* outbreaks involving 52 patients, ranging in size from 2 to 12 patients.<sup>4</sup> |
| Intensive care unit bed | An outbreak of three patients was identified, all with *Pseudomonas aeruginosa* infections and having been cared for in the same intensive care unit room over a span of five months. In response, enhanced cleaning measures were implemented for the room; no subsequent cases were identified. |
| **Transmission occurring elsewhere** |  |
| Outside facilities | Eight outbreaks, totaling 17 infections, were suspected to have origins in external facilities before transfer to our hospital. Notably, a *Proteus mirabilis* outbreak involving five patients was linked to a single skilled nursing facility over six months. The relevant external facilities were notified for further investigation and intervention. |
| Embedded, third-party chronic care facility | A 32-bed long term acute care facility operated by a third-party company is located on one floor in our hospital. There is frequent patient movement between this facility and the hospital. This unit experienced 41 infections spanning 20 outbreaks, suggesting potential transmission within the unit itself. Information about these suspected transmissions were provided to the facility's management in real-time. |
| Public Health | An outbreak of two *Pseudomonas aeruginosa* infections was identified in October 2022 without discernible commonalities.<sup>21,22</sup> However, in February 2023, the Centers for Disease Control and Prevention released details of an ongoing national outbreak linked to contaminated artificial tears, including genomic data. Subsequent analysis revealed that these two isolates were part of the national outbreak, with one patient having documented usage of the implicated eye drops just prior to infection onset.<sup>22</sup> |

### IP&C interventions and practice changes

Infection prevention initiated 134 actions which included 74 (55.2%) notification and education of staff, 25 (18.7%) enhanced cleaning efforts, 23 (17.2%) hand hygiene/personal-protective equipment compliance observations, 9 (6.7%) environmental cultures/removal of equipment, or 3 (2.2%) enhanced microbiological surveillance. IP&C initiated enhanced wound care infection prevention practices to include additional cleaning, and re-education. One intensive-care unit (ICU) began an enhanced disinfection and cleaning project that included audits using fluorescent dye. IP&C provided additional education to outpatient clinics due to suspected transmission. Due to outbreaks from shared ventilators, IP&C provided education, re-training, and enhanced cleaning to respiratory therapy. Lastly, the central sterile reprocessing team was given additional education and enhanced communication due to the nine outbreaks with suspected endoscope transmission.

### Initial detection, intervention, and subsequent infections

Among the 172 outbreaks, 121 (70.4%) had a presumed transmission route identified, which prompted targeted interventions. Of these outbreaks, 89 (73.5%) showed no further progression after two patients. There were 20 (16.5%) outbreaks that continued to increase in size, but only on identified transmission routes that had not yet been intervened upon. Only 5 (4.1%) outbreaks continually spread on the same initial transmission route after intervention. Three of the outbreaks with failed interventions were caused by VRE, one by *P. aeruginosa* in which all patients had been housed at different times in a single ICU bed (Table 2), and the last caused by *Stenotrophomonas* in a single ICU five months after the original interventions. There were 7 (5.8%) outbreaks that were detected from outside facilities that continued to spread. Overall, we observed 109/114 (95.6%) outbreaks to have no further transmission on the intervened route within our facility once IP&C intervened.

### Analysis of SNP distances and epidemiological links

Excluding *C. difficile*, there was a significant inverse relationship between pairwise SNP distance and the identification of an epidemiological link, whereby isolates with lower pairwise SNP differences were more likely linked epidemiologically (OR: 0.78; 95% CI 0.75, 0.81) (Figure S3). There were also significant associations for individual species: *Acinetobacter* (OR: 0.70; 95% CI 0.51, 0.96), *Klebsiella pneumoniae* (OR: 0.88; 95% CI 0.80, 0.97), MRSA (OR: 0.83; 95% CI 0.71, 0.99), *Pseudomonas aeruginosa* (OR: 0.77; 95% CI 0.70, 0.85), and VRE (OR: 0.78; 95% CI 0.72, 0.84).

### Economic and clinical impact analysis

A total of 62 infections were estimated to have been averted due to real-time IP&C interventions with unit-based transmission aversions representing the most infections, 36 (58.1%) (Table S2). The predominant distribution of averted infection types were respiratory and wound infections (19 and 18 infections, respectively). Considering the cost of treating by infection types, the total estimated cost aversion was $1,011,146 (Table S2). Considering the costs of a standard of care IP&C program at our institution, we estimated hospital net savings of $695,706 over the course of the two-year study period ($347,853 per year) (Table S2). This equates to a net savings of $147.3 per isolate sequenced during the study period and a 3.2-fold return on investment. PSA showed EDS-HAT as cost savings in 98% of simulations with mean savings of $660,052 (range 2.5^th^ percentile $26,383; 97.5^th^ percentile $1,769,390) and 60 infections averted (2.5^th^ percentile 22 infections averted; 97.5^th^ percentile 126 infections averted) (Figure 3).

**Figure 3.**
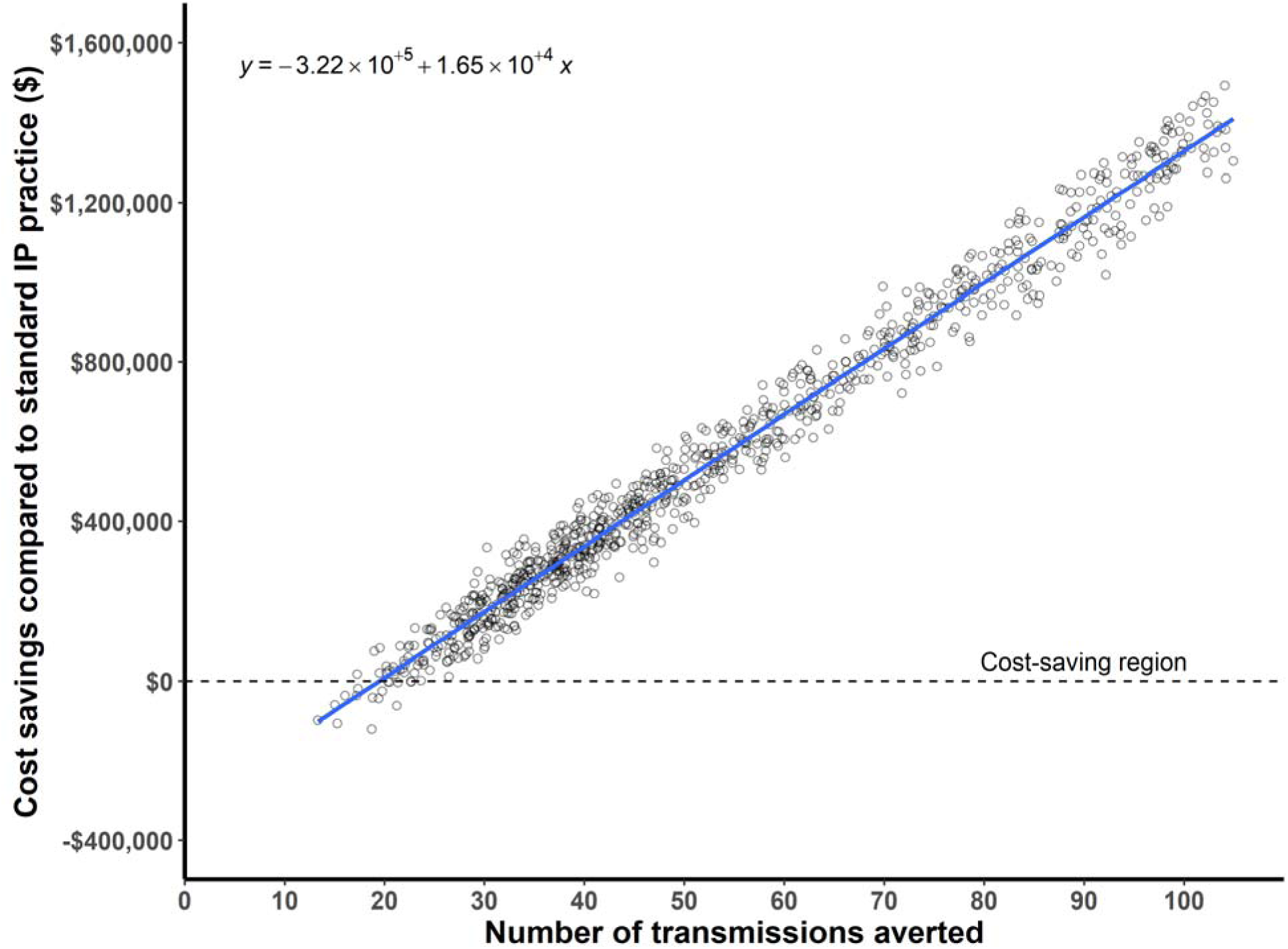
Probabilistic sensitivity analysis of cost savings. The plot shows cost savings on the y-axis and number of transmissions averted on the x-axis. Each dot represents one simulation of the model, where a random sample is drawn from the distribution of input parameters. The best fit linear model is depicted as a blue line, with the equation in the top left corner of the plot. EDSHAT was cost saving in 98% of the simulations.

## DISCUSSION

In this study, we found that genomic surveillance using WGS was successful at detecting multiple hospital outbreaks allowing for rapid intervention by our IP&C team. Due to real-time genomic surveillance, we were able to early identify outbreaks at just two cases and promptly intervene, thus preventing additional cases. Moreover, we estimated substantial net cost-savings attributed to averted infections from our interventions.

We observed that the vast majority (95.6%) of the WGS surveillance-guided IP&C interventions within an outbreak were successful at halting the subsequent spread along the same transmission route. Among the five outbreaks with noted failed interventions, three were caused by VRE. Our experience with VRE shows that transmission is driven largely by asymptomatic colonization, which is not detected by EDS-HAT.^17^ Notably, many of the outbreaks were associated with endoscopes, which are widely recognized as a common source of outbreaks within healthcare facilities.^13–15^ Traditionally, endoscope-related outbreaks often are either undetected entirely or unnoticed until multiple patients are involved.^13,15,16^ As an example, an outbreak caused by endoscopes that were contaminated by *Klebsiella pneumoniae* at our institution that occurred before we initiated WGS surveillance took 14 months to recognize and resulted in 17 cases, including 12 cases of bacteremia and five deaths.^13^ We believe that, had EDS-HAT been implemented, many of these cases would have been prevented.

Our clinical and economic modeling demonstrated an estimated 62 (21%) prevented infections and significant net savings of $695,706, with probabilistic sensitivity analysis demonstrating up to $1,769,390 in costs savings and 126 infections averted. These findings align with our previous retrospective analysis, which indicated an aversion potential ranging from 8% to 21% of transmissions. A recent study aimed at measuring the reduction of HAIs from enhanced cleaning of equipment reported a relative reduction of 34.5%, which is in line with our findings, further supporting the validity of our results.^18^

However, we believe these impact estimates may be conservative because EDS-HAT was in place for several years before this study, during which multiple outbreaks were identified and infection prevention measures were strengthened. We suspect that these measures may have reduced the number of outbreaks—a counterfactual that cannot be measured.^4,9,15,19^ For example, EDS-HAT previously identified an outbreak of VRE caused by flawed manufacturer instructions for preparation of sterile contrast material for injection into patients during interventional radiology (IR) procedures.^19^ We suspect that the outbreak would have continued or that other outbreaks could have occurred if changes in IR practice had not been implemented. Other EDS-HAT-detected outbreaks before this study led to changes in practice that could have prevented additional outbreaks during the study period. For example, after multiple wound-care-related outbreaks, a revamping of IP&C procedures appears to have successfully eliminated these incidents.

There are also other unmeasured benefits of EDS-HAT to other institutions. For instance, we identified transmissions that originated outside of our facility that were reported to the relevant IP&C teams. Ideally, a comprehensive healthcare genomic surveillance system would be implemented in the US, which would enable the identification of transmissions both within and between facilities due to patient movement.^20^ This approach would be similar to the CDC’s PulseNet network, which has transformed the detection and interruption of multi-state outbreaks caused by foodborne pathogens. Such a system could also potentially identify and track multi-institutional healthcare outbreaks linked to contaminated medications or devices more effectively.^21,22^

There are several limitations to our study. First, we did not attempt to measure a reduction in HAI rate after initiation of WGS surveillance for several reasons. We have shown that the majority of transmitted infections detected by EDS-HAT are not reportable as HAIs and therefore many of the infections prevented by EDS-HAT would not be included in an analysis of HAI rates.^23^ In addition, before and after comparisons of HAI rates and/or use of control facilities for such an analysis would be difficult to interpret given changes in IP&C practices, the stochastic nature of outbreaks, and difficulty in controlling for all confounding variables. Second, our impact analyses were highly dependent on the results of our retrospective study.^4^ The impact of EDS-HAT may have been significantly different if the actual size of the outbreaks we intervened upon were larger or smaller than what we assumed in the absence of intervention. Third, we only included the most high-risk pathogens causing clinical infections due mainly to resource constraints, which led us to miss outbreaks caused by other pathogens and transmissions from colonized individuals. Additionally, we did not include active surveillance isolates. Therefore, our analysis represents an underestimate of the potential impact of EDS-HAT if a larger array of pathogens were included. Fourth, we did not find an epidemiological link in 58 (33.7%) outbreaks. However, our analysis shows that the outbreaks or isolates without epidemiological links had higher genetic distances, suggesting that our SNP cutoffs could have been lower.

In conclusion, we have shown that real-time genomic surveillance is a feasible IP&C tool to accurately detect and stop outbreaks, while at the same time reducing costs. In essence, EDS-HAT is real-time quality improvement tool that allows for early identification of outbreaks and rapid implementation of corrective measures. The results of this study, our prior findings, and other studies suggest that genomic surveillance should be considered a standard practice in healthcare.^3–6,10,24^ As the body of evidence grows, healthcare leadership, payors, and policymakers should weigh the potential benefits of genomic surveillance for patient safety. Implementation of real-time WGS surveillance in routine IP&C practice is a paradigm shift that has the potential have substantial clinical impact and cost-savings.

## Data Availability

Illumina sequence data are available at NCBI BioProject PRJNA475751.

https://www.ncbi.nlm.nih.gov/bioproject/475751

## STUDY FUNDING

This work was funded in part by the National Institute of Allergy and Infectious Diseases, National Institutes of Health (NIH) (R01AI127472). NIH played no role in data collection, analysis, or interpretation; study design; writing of the manuscript; or decision to submit for publication.

## ACKNOWLEDGMENTS

We thank the UPMC Infection Prevention team and Paul Rhodes for his thoughtful review of the manuscript.

## Declaration of Interests

None

## Notes

### Competing Interest Statement

The authors have declared no competing interest.

### Author Declarations

IRB of the University of Pittsburgh gave ethical approval for this work.

