## Supplement Appendix for "Genomic Surveillance for Enhanced Healthcare Outbreak Detection and Control"

**Table of Contents.**

### **Supplement Methods: Clinical Impact Modeling Rationale and Assumptions.**

Genomic surveillance directs Infection Prevention and Control (IP&C) interventions to halt outbreaks on specific units or equipment identified as the suspected causative agents for transmission. These interventions enhance the baseline infection prevention practices on the affected units or equipment, effectively halting additional spread. Infected patients can move to new units where no directed IP&C interventions may be in place that enhance the baseline IP&C practices. This can lead to subsequent spread on those units, which are not yet addressed by genomic surveillance interventions (**Figure S1**). Additional spread in new units or procedures where transmission has not yet been detected and intervened is not a failure of genomic surveillance but rather a limitation of the baseline IP&C practices. Therefore, analyzing the overall outbreak size or number of outbreaks may be misleading. The effectiveness of genomic surveillance may be best evaluated by studying the downstream spread on an intervened route once interventions are implemented. This can be modeled against what typically occurs in genomic surveillance periods where no intervention is made (**Figure S2**).

We also conducted a probabilistic sensitivity analysis (PSA) to evaluate how uncertainty in input parameters affects the clinical and economic outcomes of EDS-HAT. For this analysis, we sampled 1,000 random draws of input parameters from their respective distributions (e.g., beta distribution for proportions, gamma distribution for costs etc.) (**Table S1**). We then calculated the proportion of simulations in which EDS-HAT was more effective and less costly than the standard IP&C practice. **Figure 3** shows that EDS-HAT was a cost-saving and more effective program in 98% of the simulations.

### **Supplement Methods: Bacterial Species Included for Sequencing**

Patients with clinical cultures positive for *Acinetobacter* species, *Burkholderia* *cepacia*, *Citrobacter* species, *Clostridioides* *difficile*, *Enterobacter* species, Extended-spectrum beta-lactamase (ESBL) *Escherichia* *coli*, ESBL *Klebsiella* species, *Legionella* species, methicillin-resistant *Staphylococcus* *aureus*, *Proteus* *mirabilis*, *Providencia* species, *Pseudomonas* species, *Serratia* species, *Stenotrophomonas* *maltophilia*, or vancomycin-resistant *Enterococcus* species were included if the patient had been in the hospital for ≥3 days or had a recent UPMC healthcare exposure in the prior 30 days. For *Clostridioides* *difficile*, culture-independent diagnostic test-positive stool specimens were cultured for the organism. Active surveillance cultures were not included.

**Table S1. Input parameters for clinical and economic impact analysis**

| **Parameter** | **Base case value** | **Distribution** | **95% CI^a^** | **Source^b^** |
| --- | --- | --- | --- | --- |
| Distribution of real time outbreaks by transmission route | | | | |
| Instrument | 12 | Not varied^g^ |  |  |
| Procedure | 11 | Not varied^g^ |  |  |
| Provider | 9 | Not varied^g^ |  |  |
| Unit | 102 | Not varied^g^ |  |  |
| Proportion of outbreaks with size > 2 by transmission route – Retrospective surveillance | | | | |
| Instrument | 67% | Beta | [28%, 95%] |  |
| Procedure | 20% | Beta | [3%, 48%] |  |
| Provider | 23% | Beta | [5%, 48%] |  |
| Unit | 23% | Beta | [14%, 35%] |  |
| Proportion of outbreaks with size > 2 by transmission route – Real time surveillance | | | | |
| Instrument | 0% | Beta | [0%, 0%] |  |
| Procedure | 0% | Beta | [0%, 0%] |  |
| Provider | 0% | Beta | [0%, 0%] |  |
| Unit | 11% | Beta | [6%, 17%] |  |
| Average number of patients additionally infected beyond first 2 patients – Retrospective surveillance | | | | |
| Instrument | 1.25 | Empirical^c^ | [1, 2] |  |
| Procedure | 6.00 | Empirical^c^ | [6, 6] |  |
| Provider | 1.33 | Empirical^c^ | [1, 2] |  |
| Unit | 2.00 | Empirical^c^ | [1, 4] |  |
| Average number of patients additionally infected beyond first 2 patients – Real time surveillance | | | | |
| Instrument | 0 | Empirical^c^ | [0, 0] |  |
| Procedure | 0 | Empirical^c^ | [0, 0] |  |
| Provider | 0 | Empirical^c^ | [0, 0] |  |
| Unit | 1.09 | Empirical^c^ | [1, 2] |  |
| Attributable mortality risk due to infection | | | | |
| Wound | 0.028 | Beta | [0.028, 0.029] | ^1^ |
| Urinary tract | 0.023 | Beta | [0.023, 0.024] | ^1^ |
| Bacteremia | 0.123 | Beta | [0.122, 0.125] | ^1^ |
| *Clostridioides difficile* | 0.030 | Beta | [0.029, 0.031] | ^2^ |
| Pneumonia | 0.144 | Beta | [0.142, 0.145] | ^1^ |
| Cost of treating infection^d,e,f^ |  |  |  |  |
| Wound (T81.4XXA) | $17,958 | Gamma | [$17,553 $18,367] | ^3,4^ |
| Urinary tract (N39.0) | $8,270 | Gamma | [$8,148 $8,394] | ^3,4^ |
| Bacteremia (R78.81) | $13,939 | Gamma | [$13,142 $14,758] | ^3,4^ |
| *C. difficile* infection (A04.7) | $11,066 | Gamma | [$10,739 $11,398] | ^3,4^ |
| Pneumonia (J15.0) | $22,324 | Gamma | [$19,357 $25,499] | ^3,4^ |
| Number of IP professionals in team |  |  |  |  |
| EDS-HAT | 8 | Not varied^g^ |  | Unpublished |
| SoC | 8 | Not varied^g^ |  | Unpublished |
| Annual salary of an IP professional^e^ | $101,272 | Gamma | [$100,009 $102,542] | ^5^ |
| % of time spent on IP activities |  |  |  |  |
| EDS-HAT | 10% | Beta | [8%, 12%] | Unpublished |
| SoC | 10% | Beta | [8%, 12%] | Unpublished |
| Unit cost of sequencing |  |  |  |  |
| EDS-HAT | $80 | Gamma | [$65, $96] | ^6^ |
| SoC | $240 | Gamma | [$195, $289] | Unpublished |

a The 95% CI column represents the confidence interval for parameters except for ‘% of time spent on IP activities and unit cost of sequencing, where the column represents the uncertainty range, estimated assuming a 10% deviation around the mean value.

b Unpublished refers to the internal data of the UPMC Presbyterian Hospital.

c Empirical distribution is based on the cluster-level data from retrospective and prospective period.

d It was assumed that all positive cultures from wound, urine, blood, stool and respiratory represented infections.

e All costs were adjusted to 2023 using medical component of Consumer Price Index (CPI) obtained from Bureau of Labor Statistics.

f ICD 10 codes are provided in parenthesis

g Some parameters such as the total number of outbreaks observed in the study period and number of IP professionals in IP team were considered fixed and hence not varied in probabilistic sensitivity analysis.

Abbreviations: EDS-HAT, Enhanced Detection System for Healthcare-Associated Transmission; ICD, International Classification of Diseases; IP, Infection Prevention; SoC, Standard of Care

### **Table S2**. **Model results of estimated number of averted infections and associated costs**

|  | **Standard of care (A)** | **EDS-HAT (B)** | **Change (A-B)** |
| --- | --- | --- | --- |
| **Costs Estimated, Averted** |  | | |
| Whole genome sequencing costs | $62,400 | $377,840 | -($315,440) |
| Infection treatment costs | $4,706,301 | $3,695,155 | $1,011,146 |
| IP&C program costs | $162,035 | $162,035 | $0 |
| Total costs | $4,930,736 | $4,235,030 | **$695,706 saved** |
| **Infections and Deaths Estimated, Averted** |  | | |
| Respiratory | 88 | 69 | 19 |
| Wound | 84 | 66 | 18 |
| Blood | 45 | 35 | 10 |
| Urine | 58 | 46 | 13 |
| Stool / *C. difficile* | 11 | 9 | 2 |
| **Total number of infections** | 287 | 225 | **62 averted** |
| **Total number of expected deaths** | 22.2 | 17.4 | **4.8 saved** |

The economic analysis covers the two-year period i.e., 2021-2023. Costs are reported in 2023 dollars.

Abbreviations: EDS-HAT, Enhanced Detection System for Healthcare-Associated Transmission; IP&C, Infection prevention and control

### **Table S3. Outbreak size distribution by species.**

|  | **Outbreaks by Size (Patients)** | | | | | | | | | | | | |
| --- | --- | --- | --- | --- | --- | --- | --- | --- | --- | --- | --- | --- | --- |
| **Organism** | **2** | **3** | **4** | **5** | **6** | **7** |  | **9** | **10** | **11** | **.** | **16** | **Average Outbreak Size** |
| *Acinetobacter* species | 7 |  | 2 |  |  |  |  |  |  |  |  |  | 2.4 |
| *Citrobacter* species | 2 |  |  |  |  |  |  |  |  |  |  |  | 2.0 |
| *Clostridioides* *difficile* | 5 | 1 |  |  | 1 |  |  |  |  |  |  |  | 2.7 |
| *Enterobacter* *cloacae* | 3 | 1 |  |  |  |  |  |  |  |  |  |  | 2.3 |
| *Escherichia* *coli*, ESBL producing | 5 | 4 |  |  |  |  |  |  |  |  |  |  | 2.4 |
| *Klebsiella* *oxytoca* | 2 |  |  |  |  |  |  |  |  |  |  |  | 2.0 |
| *Klebsiella* *pneumoniae*, ESBL producing | 13 | 4 |  |  | 1 |  |  |  |  | 1 |  |  | 2.9 |
| Methicillin-resistant *Staphylococcus* *aureus* | 17 | 6 |  |  |  |  |  |  |  |  |  |  | 2.3 |
| *Proteus* *mirabilis* | 8 |  |  | 1 |  |  |  |  |  |  |  |  | 2.3 |
| *Providencia* species | 1 |  |  |  |  |  |  |  |  |  |  |  | 2.0 |
| *Pseudomonas* *aeruginosa* | 31 | 7 | 2 | 1 | 1 | 1 |  |  |  |  |  |  | 2.5 |
| *Pseudomonas* species, not aeruginosa | 1 |  |  |  |  |  |  |  |  |  |  |  | 2.0 |
| *Serratia* *marcescens* | 6 | 1 |  |  |  |  |  |  |  |  |  |  | 2.1 |
| *Stenotrophomonas* *maltophilia* | 5 | 1 |  |  |  |  |  |  |  |  |  |  | 2.2 |
| Vancomycin-resistant *Enterococcus* *faecalis* | 13 | 5 | 3 | 2 |  | 2 |  |  | 1 | 2 |  | 1 | 4.3 |
| **TOTAL** | **120** | **30** | **7** | **4** | **3** | **3** |  |  | **1** | **3** |  | **1** | **2.7** |

ESBL: Extended-spectrum beta-lactamase

**Figure S1. Hypothetical outbreak scenario.** This outbreak illustrates how genomic surveillance effectively directs Infection Prevention and Control (IP&C) interventions to halt outbreaks on specific units yet the outbreak can still increase in overall size. Figure made with BioRender.

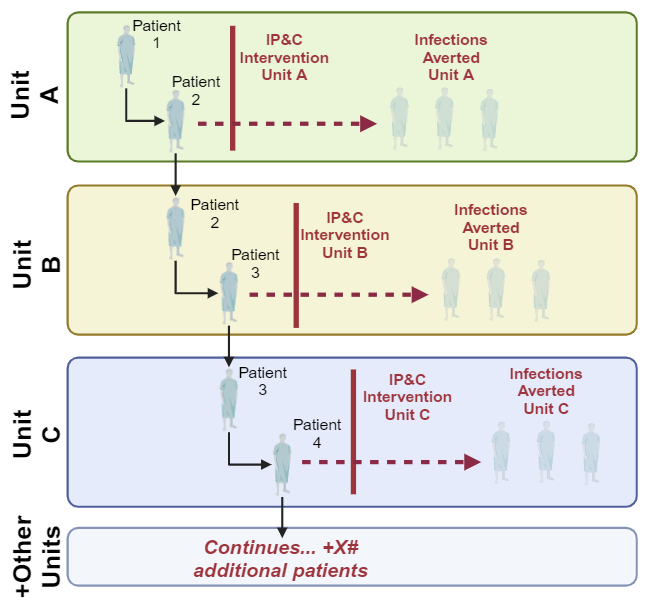

**Figure S2. Hypothetical data scenario.** Hypothetical data to approximate infections averted using genomic surveillance. Outbreaks with patients ≥3 patients on the same transmission route are utilized. Retrospective genomic surveillance without any Infection Prevention and Control (IP&C) interventions is applied on the number of outbreaks detected by transmission route type in prospective genomic surveillance to estimate the number of infections expected. The observed outbreak dynamics are then subtracted from the expected number of infections. Figure made with BioRender.

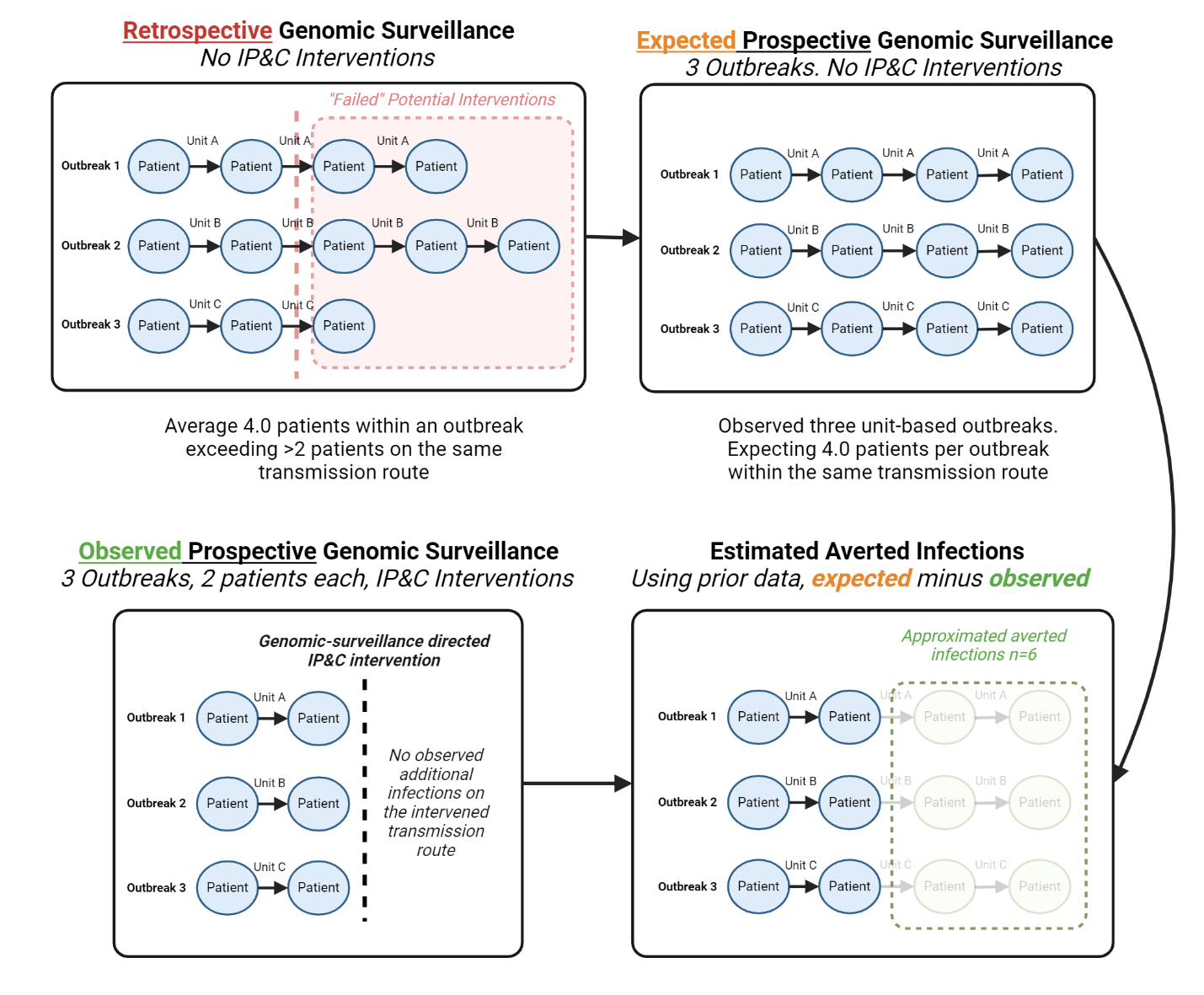

**Figure S3. Isolate pairs with an identified transmission route, by single pairwise nucleotide polymorphism (SNP) distance.**

Percent Isolates With Transmission Route Identified

Number of Pairs

| Pairwise SNP Distance | % Isolates with Epi Link | % Isolates without Epi Link | Number of Pairs |
| --- | --- | --- | --- |
| 0 | 75.6098 | 24.3902 | 41 |
| 1 | 81.1321 | 18.8679 | 53 |
| 2 | 51.2195 | 48.7805 | 41 |
| 3 | 66.6667 | 33.3333 | 39 |
| 4 | 53.8462 | 46.1538 | 39 |
| 5 | 38.2979 | 61.7021 | 47 |
| 6 | 35.2941 | 64.7059 | 34 |
| 7 | 21.4286 | 78.5714 | 28 |
| 8 | 31.5789 | 68.4211 | 38 |
| 9 | 17.0732 | 82.9268 | 41 |
| 10 | 21.2121 | 78.7879 | 33 |
| 11 | 21.6216 | 78.3784 | 37 |
| 12 | 10.4167 | 89.5833 | 48 |
| 13 | 4.44444 | 95.5556 | 45 |
| 14 | 11.9048 | 88.0952 | 42 |
| 15 | 4.16667 | 95.8333 | 24 |
| 16 | 8.33333 | 91.6667 | 24 |
| 17 | 11.7647 | 88.2353 | 17 |
| 18 | 6.25 | 93.75 | 16 |
| 19 | 0 | 100 | 9 |
| ≥20 | 2.94118 | 97.0588 | 34 |

**References**.
